# Contrastive Learning Model for Wearable-based Ataxia Assessment

**DOI:** 10.1101/2025.02.28.25323114

**Authors:** Juhyeon Lee, Brandon Oubre, Jean-Francois Daneault, Christopher D. Stephen, Jeremy D. Schmahmann, Anoopum S. Gupta, Sunghoon Ivan Lee

**Affiliations:** Manning College of Information and Computer Sciences, University of Massachusetts Amherst, MA, USA; Department of Computer Science, University of Alabama at Birmingham, AL, USA; Department of Neurology, Massachusetts General Hospital, Harvard Medical School, MA, USA; Department of Rehabilitation and Movement Sciences, Rutgers University, NJ, USA; Department of Neurology and the Ataxia Center, Massachusetts General Hospital, Harvard Medical School, MA, USA

**Keywords:** Cerebellar Ataxia, Contrastive Learning, Motor Severity Assessment, Wearable Sensors

## Abstract

**Objective:** Frequent and objective assessment of ataxia severity is essential for tracking disease progression and evaluating the effectiveness of potential treatments. Wearable-based assessments have emerged as a promising solution. However, existing methods rely on inertial data features directly correlated with subjective and coarse clinician-evaluated rating scales, which serve as imperfect gold standards. This approach may introduce biases and restrict flexibility in feature design. To address these limitations, this study introduces a novel contrastive learning-based model that leverages motor severity differences in wearable inertial data to learn relevant features.

**Methods:** The model was trained on inertial data collected from 87 individuals with diagnostically heterogeneous ataxias and 44 healthy participants performing the finger-to-nose task. A pairwise contrastive loss function was proposed to learn representations capturing relative differences in ataxia severity, which were evaluated through downstream regression and classification tasks.

**Results:** The learned features demonstrated strong cross-sectional (r = 0.84) and longitudinal (r = 0.68) associations with clinical scores and robust measurement reliability (intraclass correlation coefficient = 0.96). Additionally, the model exhibited strong known-group validity, distinguishing between ataxia and healthy phenotypes with an area under the receiver operating characteristic curve of 0.95.

**Conclusion:** The proposed contrastive model captures robust representations of disease severity with reduced reliance on clinical scales, outperforming state-of-the-art methods that derive features directly from clinical scores.

**Significance:** Combining wearable sensors with contrastive learning enables a more objective, scalable, and frequent method for assessing ataxia severity, with the potential to enhance patient monitoring and improve clinical trial efficiency.

## I. Introduction

ATAXIAS are a group of neurologic disorders characterized by impaired coordination and balance, resulting from diverse underlying diseases [1]. Many ataxias are progressive, leading to worsening motor impairments that significantly affect the ability to perform daily activities, thereby reducing the quality of life and independence of those affected [2], [3]. Frequent and objective assessments of ataxia severity are essential for tracking disease progression and providing appropriate medical interventions [3]. Furthermore, the lack of disease-modifying treatments for many ataxias highlights the importance of developing improved outcome measures to facilitate ongoing clinical trials [4], [5]. These clinical trials would benefit from frequent and sensitive endpoints to accurately evaluate drug efficacy and expedite the development process of new therapies [6], [7].

Currently, ataxia severity is assessed by neurologists using clinical rating scales, such as the Brief Ataxia Rating Scale (BARS) [8] and the Scale for the Assessment and Rating of Ataxia (SARA) [9]. These assessments rely on clinicians’ visual observations of patients performing predefined motor tasks, which typically necessitate in-person visits with specialized neurologists. This requirement limits the frequency of assessments and cannot support continuous monitoring. Moreover, the subjective nature and coarse numerical structure of these scales further limit their precision, reducing their sensitivity to subtle changes in disease progression [10], [11].

Recent studies have demonstrated the potential of wearable sensors and machine learning techniques to support the frequent and objective assessment of ataxia severity [10], [12]–[19]. Most previous approaches have relied on manually extracting features from wearable sensor data collected during motor tasks, which are then used to train machine learning models for ataxia severity assessment. However, these methods often constrain feature design and selection to established knowledge of ataxia symptoms and their direct correlations with clinical scores, limiting flexibility and introducing potential biases.

To overcome these shortcomings, several studies have applied deep learning models to automate the feature learning process, though primarily in the context of other neurological diseases rather than ataxia [20], [21]. While promising, these deep learning models, like conventional machine learning approaches, are trained to learn features by using clinician-evaluated rating scales as the reference target variable, which are known to have considerable measurement errors [22]. Directly using clinical scales as the learning objective can cause models to internalize inherent errors and systematic biases. Ideally, wearable-based motor assessments should aim to quantify the *true severity differences* in motor function over time and across individuals, a latent characteristic, rather than replicating clinical scores.

In this study, we introduce a novel supervised contrastive learning-based approach to capture motor representations of ataxia severity. Supervised contrastive learning leverages labeled data to form positive and negative pairs [23]. The model is then trained to differentiate between similar (positive) and dissimilar (negative) data pairs, learning meaningful, generalizable representations of the data that are robust to noisy labels [24], [25]. Our algorithm, therefore, is specifically designed to learn features that produce similar values for patients with comparable clinician-evaluated severity and distinct values for those with differing severity, without directly fitting to clinician-evaluated scores, as illustrated in Fig. 1. For example, consider two patients with the same underlying motor severity but slightly different clinical scores due to measurement errors. Under the hypothesis that wearable sensor data provide more consistent and objective information about motor severity, patients’ motor task performance—as captured by inertial sensor data—is likely to be much more similar compared to patients with substantially different motor severity and clinician-evaluated scores. This pairwise approach enables the model to identify precise and sensitive patterns, even in the presence of measurement errors in clinician-evaluated reference scores.

**Fig. 1.**
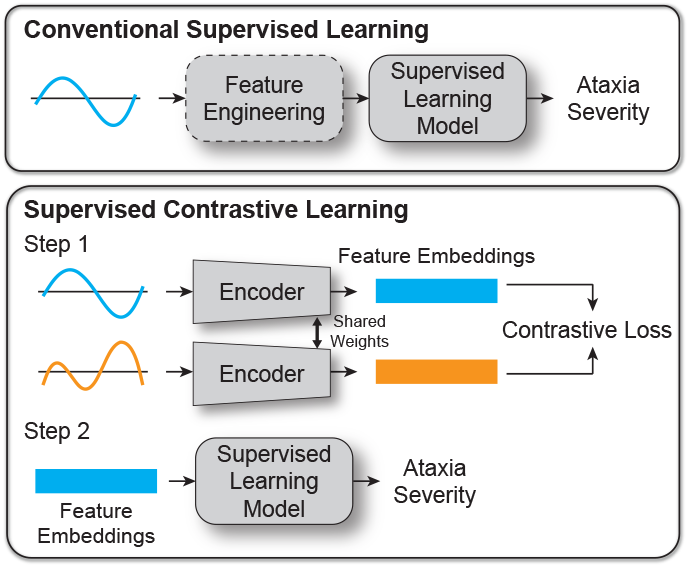
Conventional supervised learning vs. supervised contrastive learning approaches for wearable-based assessment of ataxia severity. The conventional supervised learning method relies on either manual feature engineering or learning features that directly correlate with clinician-evaluated motor assessments, which are imperfect gold standards. In contrast, the proposed supervised contrastive learning approach uses an encoder to learn feature embeddings via a contrastive loss function, capturing implicit differences inherent in the data. For both approaches, the extracted features are aggregated into a single metric representing the patient’s motor severity using a supervised machine learning model, with cost functions specifically designed to optimize validation criteria for biomedical measurements, such as reliability, convergent validity, and/or responsiveness to disease progression.

We demonstrate the proposed contrastive learning model by training it on inertial data collected from 131 study participants (87 individuals with ataxia and 44 healthy controls) using two wrist-worn sensors during the finger-to-nose task. The learned representations were evaluated in downstream tasks, showing strong convergent validity with clinician-scored severity, responsiveness, and known-groups validity. The findings suggest that contrastive learning-based approaches have the potential to provide a more objective and sensitive method for monitoring disease progression and evaluating the effectiveness of treatments. The proposed machine learning framework could also be adapted to other sensing devices and diseases characterized by motor impairments, broadening its applicability. To facilitate further research, the source code and trained models are publicly available on GitHub (https://github.com/juhyeonlee/AtaxiaSeverity-Contrastive-Model).

## II. Methods

### A. Data Collection

A total of 87 individuals with clinically and/or genetically diagnosed ataxias and 44 neurologically healthy individuals were recruited from Massachusetts General Hospital between September 2017 and March 2020. Participants met the following inclusion criteria: 1) age between 2 and 90 years, 2) either a clinical diagnosis of ataxia or being neurologically healthy, and 3) the ability to perform the instrumented finger-to-nose task. Demographic details are provided in Table I. The study protocol was approved by the Institutional Review Board at Massachusetts General Hospital (IRB #: 2016P001048), and all participants provided written informed consent or assent.

**TABLE I.** Demographic Characteristics OF Study Participants.

|  | Ataxia | Healthy |
| --- | --- | --- |
| $N$ | 87 | 44 |
| Age (years) | $54.3 \pm 18.5$<br>(range: 5-78) | $32.9 \pm 19.6$<br>(range: 9-86) |
| Sex | 46 male,<br>41 female | 22 male,<br>22 female |
| Handedness | 77 right,<br>9 left<br>1 ambidextrous | 40 right,<br>4 left |
| Motor Severity<br>(total BARS scores) | $10.5 \pm 5.4$<br>(range: 0-24) | — |
| Arm Severity<br>(BARS Finger-to-nose<br>subscores) | $1.9 \pm 1.4$<br>(range: 0-6) | — |
| Clinical Diagnoses | 4 SCA-1, 2 SCA-2,<br>11 SCA-3, 7 SCA-6,<br>10 other SCA, 7 A-T, 3 FA,<br>7 MSA-C, 1 PSP-C, 3 HSP,<br>4 AIA, 1 BD, 1 HE, 2 EA,<br>2 ARCA-1, 1 ARCA-3,<br>1 CH, 3 DN, 1 SA,<br>1 FXTAS, 1 GHS,<br>1 LCHND, 1 SRA, 3 TA,<br>2 SAOA, 5 SAOAN,<br>2 ADCA | — |

Mean ± Standard deviation; BARS: Brief Ataxia Rating Scale; SCA: spinocerebellar ataxia; A-T: Ataxia-Telangiectasia; FA: Friedreich's Ataxia; MSA-C: multiple system atrophy, cerebellar-type; PSP-C: progressive supranuclear palsy, cerebellar-dominant; HSP: hereditary spastic paraplegia; AIA: autoimmune-related ataxia with undefined cause; BD: Behcet's Disease; HE: Hashimoto's Encephalopathy; EA: episodic ataxia; ARCA: autosomal recessive cerebellar ataxia; CH: cerebellar hypoplasia; DN: Downbeat Nystagmus with mild ataxia; SA: sensory ataxia; FXTAS: Fragile XAssociated Tremor/Ataxia Syndrome; GHS: Gordon Holmes' Syndrome; LCHND: LCH-related neurodegeneration; SRA: stroke-related ataxia; TA: transient ataxia, later resolved; SAOA: sporadic adult-onset ataxia; SAOAN: sporadic adult-onset ataxia with neuropathy; ADCA: autosomal dominant cerebellar ataxia with unidentified genetic cause;

Participants performed an instrumented finger-to-nose task while wearing nine-axis Inertial Measurement Units (IMU; Opal, APDM Wearable Technologies) on both wrists [12]. Conventionally, the finger-to-nose task requires participants to move their finger between their nose and a clinician’s finger as quickly and accurately as they can. In this study, a 12.9inch tablet device (iPad Pro, Apple Inc.) was used to display a reaching target in place of the clinician’s finger, as described in prior work [12]. Participants performed the continuous fingerto-nose task for 40 *s* with each arm. Fourteen individuals with ataxia and five healthy individuals participated in two data collection sessions at different time points, while one individual with ataxia participated in four sessions. All sessions took place on separate visits, with an average interval of 299.0 ± 107.1 days (range: 126–452 days) between sessions.

Motor impairment severity in ataxia participants was evaluated by a neurologist using the Brief Ataxia Rating Scale (BARS), which ranges from 0 to 30 in half-point increments, with higher scores indicating greater motor impairment. The total BARS score is calculated by summing subscores from assessments of finger-to-nose, knee-tibia, gait, speech, and oculomotor performance. Healthy participants were assigned a score of 0, representing no motor impairment, without administering the assessment.

### B. Preprocessing of Inertial Data

Raw accelerometer, gyroscope, and magnetometer data were preprocessed to reduce noise and generate linear velocity time-series before being applied to the contrastive model. The inertial data, sampled at 128 *Hz*, were processed to generate gravity-free acceleration time-series in a global coordinate frame using a manufacturer-provided sensor fusion algorithm [26]. While the *z*-axis of the gravity-free acceleration was aligned opposite to gravity, the *x*-*y* plane was aligned to Earth’s magnetic north pole. To remove non-human-generated, high-frequency noise, the gravity-free acceleration time-series were low-pass filtered using a fifth-order Butterworth filter with a cut-off frequency of 20 *Hz* [12]. The filtered acceleration data were then integrated to yield velocity time-series, which were further band-pass filtered using cut-off frequencies of 0.1 *Hz* and 20 *Hz* to attenuate integration drift and noise [12], [27]. The start and end of each movement were determined through visual inspection of the filtered velocity time-series, with movement onset defined as the point where the signal magnitude increased and was followed by clear, repetitive patterns [12]. The resulting trimmed velocity timeseries were segmented using a sliding window with a fixed segment length *L* for model input. Velocity time-series were used instead of acceleration, as previous studies have demonstrated that velocity profiles effectively capture the kinematic characteristics of ataxic movements [12], [13].

### C. Ataxia Severity Contrastive model

The overall model pipeline is illustrated in Fig. 2. Velocity segments, each with a length *L*, were fed into the model as inputs. The model encoded representations (i.e., the feature embeddings) from each velocity segment using the proposed pairwise contrastive loss. The encoder architecture consisted of a linear projection layer, a random masking layer, twenty dilated convolution layers, and max-pooling layers, which had proven effective for various time-series data in prior work [28]. The proposed loss function was inspired by the generalized InfoNCE loss, originally designed to contrast inputs versus augmentations in a self-supervised manner [29]. To capture fine-grained ataxia severity, our approach extended InfoNCE loss to a supervised framework, contrasting time-series segments from individuals with varying ataxia severity using clinician-evaluated ratings (i.e., BARS) [30]. This method allows the model to capture relative differences in ataxia severity within and across individuals.

**Fig. 2.**
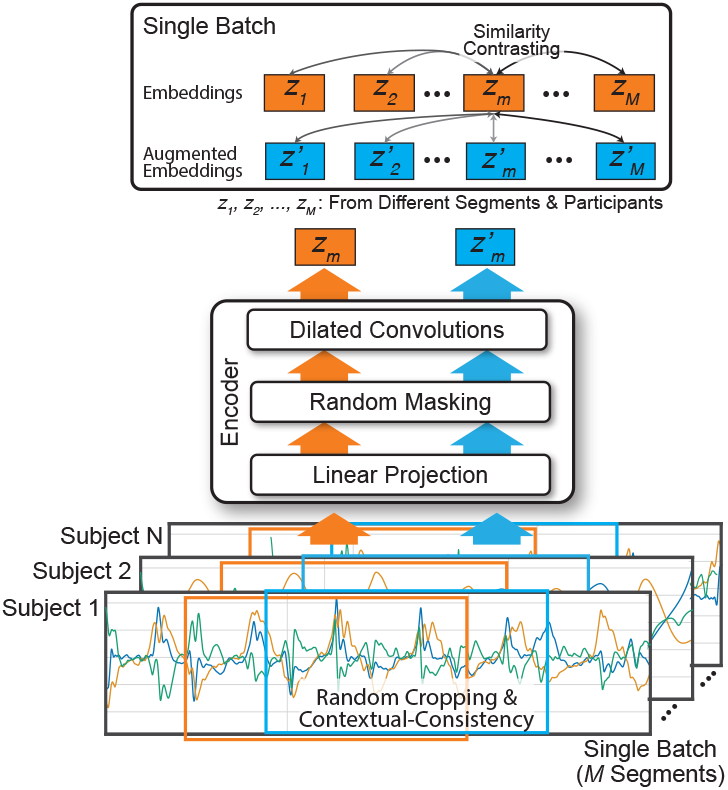
Overview of the contrastive learning model for ataxia severity assessment based on wrist-worn sensor velocity segments. The model employs an encoder with linear projection, random masking, and dilated convolutions to generate feature embeddings. Random cropping and contextual consistency augmentations are used to create feature embeddings (***z***_***i***_) and augmented embeddings 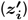 from the same segments. Similarity contrasting is then applied between embeddings from different participants’ segments to enforce similarity proportional to ataxia severity.

Formally, let the *m*^*th*^ segment of a participant’s velocity time-series be **x**_*m*_ ∈ ℝ^*L×*3^. Each segment was fed into the encoder *f* (·) : ℝ^*L×*3^ → R^*D*^ to generate the feature embedding **z**_*m*_ = *f* (**x**_*m*_; *θ*) ∈ ℝ^*D*^ for learned parameters *θ*. To enhance the learning process, we generated augmented feature embedding **z**^*′*^_*m*_ ∈ ℝ^*D*^ using random masking and contextual-consistent random cropping as described in prior work [28]. Thus, a single batch consisted of two sets of feature embeddings, 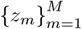 and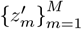, resulting in a total of 2 × *M* embeddings. This combined set of feature embeddings can be denoted as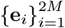, where **e**_*i*_ = *z*_*i*_ for *i* ≤ *M* and 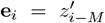 for *i > M*. The corresponding set of clinician-evaluated BARS scores is represented as 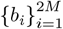. The learned encoder parameters *θ* were iteratively optimized to enable the feature embeddings to capture the severity of ataxia by minimizing the proposed loss function ℒ, shown in (1), for each batch. ℒ imposes the similarity of learned representations proportional to the similarities in ataxia severity. In other words, the encoder was trained to increase the similarity between feature embeddings for segments with similar BARS scores and decrease the similarity between feature embeddings for segments with large differences in BARS scores. ℒ contrasted each feature embedding with all other embeddings, both original and augmented, except itself. In (1), sim(·, ·) represents the dot-product similarity measure between two feature embeddings.

To capture robust representations across different temporal scales, a hierarchical loss was applied using max-pooling layers during training, as described by Yue *et al*. [28]. The max-pooling layer used a kernel size of 2, applied iteratively until the feature embedding dimensions were reduced to a single 320-dimensional vector. The loss function (1) was computed at each temporal level and summed to form the total loss. During testing, max-pooling was applied once to produce the final 320-dimensional feature embedding.

The final feature embeddings for downstream tasks were aggregated by averaging the values of multiple 320-dimensional feature embeddings generated from all velocity segments pertaining to each participant’s session. This resulted in a single 320-dimensional vector per session. These vectors were paired with their respective BARS scores for training and testing downstream tasks.

### D. Contrastive Learning Model Training

We trained contrastive models with varying input segment lengths *L* to observe its impact on model performance and learned representations. The segment length *L* was varied, starting from 0.5 *s* and then increasing from 1 *s* to 20 *s* in increments of 1 *s*. This variation was chosen because subsequent analysis of the reliability of the wearable-based motor assessment (Section II-E) required segments to be at most half the duration of the finger-to-nose task (i.e., 40 *s*). For the training set, input segments were created using a sliding window with an overlap of *L/*2 to augment the training data. In contrast, non-overlapping windows were used for the testing set.

Prior work using wearable sensors to analyze the finger-to-nose task required the identification of a participant-specific,

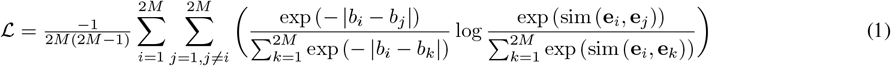

3D coordinate frame, such as one aligned with the participant’s facing direction [12]. However, to eliminate this additional processing and enhance the model’s robustness to orientation variability, we applied data augmentation during training by randomly rotating each velocity segment within the transverse plane.

The model was optimized using the Adam optimizer with decoupled weight decay [31], a batch size of 32, and a learning rate of 10^*−*3^. The number of training epochs was 100 for a segment length of 1 second, 50 for 0.5 seconds, and 200 for other segment lengths. The number of epochs was determined based on the point at which the training loss plateaued.

### E. Evaluation of the Representations Learned by the Contrastive Model

To evaluate the effectiveness of the learned feature embeddings in capturing ataxia severity, we conducted two downstream tasks: a classification task to assess known-group validity and a regression task to assess convergent validity and responsiveness. Both tasks were performed using five-subject-fold cross-validation, ensuring that no participant appeared in both the contrastive learning and the downstream tasks to ensure generalizability to unseen participants.

First, a Support Vector Classifier (SVC) was trained to distinguish between healthy individuals and those with ataxia. This task evaluated whether the learned representations encoded sufficient information to differentiate between groups known to differ on the construct of interest (i.e., motor severity), analogous to known-group validity. Classification performance was evaluated using the Area Under the Receiver Operating Characteristic Curve (AUC-ROC).

Second, we developed a wearable-based assessment of ataxia motor severity by training a Support Vector Regression (SVR) model. This regression task aggregated the learned features into a single quantitative measure representing patients’ motor severity. Since convergent validity is a key validation criterion for motor assessments, the SVR was trained to maximize its cross-sectional association with clinician-evaluated motor severity (i.e., BARS scores). Cross-sectional association was evaluated using Pearson’s correlation. Moreover, we evaluated the responsiveness of the wearable-based assessment by calculating Pearson’s correlation between its changes and corresponding changes in their BARS scores among participants with multiple clinical visits. This longitudinal correlation analysis quantified whether temporal changes in the wearable-based estimates aligned with changes observed in clinician-rated motor severity, thereby indicating the model’s responsiveness to disease progression.

We further assessed the reliability of the wearable-based motor assessments through two approaches. First, we evaluated the agreement of motor severity assessments based on feature embeddings derived from the first half versus the second half of the motor task. Feature embeddings from the first and second halves of the time-series were separately processed to generate motor severity predictions using the trained SVR model, and their agreement was measured. Second, we examined the agreement in motor severity assessments between the left and right wrists. In this analysis, feature embeddings from each wrist were separately processed to generate predictions with the trained SVR model. Reliability was quantified using a two-way mixed effects, absolute agreement, single rater Intraclass Correlation Coefficient (computationally equivalent to ICC(2,1)) [32].

To visualize the feature embeddings, we projected them into a two-dimensional space using Principal Component Analysis (PCA). PCA was fitted to the averaged feature embeddings derived from each participant’s session in the training set of each cross-validation fold. The learned transformation was then applied to the corresponding test set of that fold to ensure a consistent projection across training and test data.

## III. Results

### A. Task Performance across Segment Lengths

Table II summarizes the regression and classification results for various input segment lengths. For the classification task, the AUC-ROC values ranged from 0.92 to 0.97, indicating outstanding discriminative performance across different segment lengths [33]. For the regression task, the correlation coefficients ranged from 0.82 to 0.88, demonstrating strong convergent validity [34]. Significant longitudinal correlations were observed for most segment lengths, ranging from 0.49 to 0.68, with the exceptions of the 1 s, 4 s and 18 s segments, which had correlation coefficients of 0.23, 0.45, and 0.44, respectively. Reliability assessments revealed high ICC values, with agreements within a single time-series ranging from 0.92 to 0.97, indicating good to excellent reliability [32].

**TABLE II.** Downstream TASK PERFORMANCE ACROSS DIFFERENT SEGMENT LENGTHS.

| Segment Length (s) | AUC-ROC (Healthy vs. Ataxia) | Cross-sectional correlation ($N = 131$) | | ICC(2, 1) Left- vs. Right-arm | ICC(2, 1) First- vs. Second-half time-series | Longitudinal correlation ($N = 15$) | |
| --- | --- | --- | --- | --- | --- | --- | --- |
|  |  | r | p |  |  | r | p |
| 0.5 | 0.94 | 0.84 | < 0.001* | 0.85 [0.8 0.89] | 0.95 [0.93 0.96] | 0.51 | 0.038* |
| 1.0 | 0.92 | 0.84 | < 0.001* | 0.84 [0.79 0.88] | 0.95 [0.93 0.96] | 0.23 | 0.371 |
| 2.0 | 0.94 | 0.84 | < 0.001* | 0.88 [0.84 0.91] | 0.96 [0.94 0.97] | 0.64 | 0.006* |
| 3.0 | 0.94 | 0.84 | < 0.001* | 0.89 [0.86 0.92] | 0.95 [0.93 0.96] | 0.49 | 0.048* |
| 4.0 | 0.95 | 0.84 | < 0.001* | 0.89 [0.85 0.92] | 0.96 [0.94 0.97] | 0.45 | 0.069 |
| 5.0 | 0.93 | 0.82 | < 0.001* | 0.88 [0.84 0.91] | 0.96 [0.94 0.97] | 0.66 | 0.004* |
| 6.0 | 0.94 | 0.82 | < 0.001* | 0.88 [0.84 0.91] | 0.95 [0.93 0.96] | 0.57 | 0.016* |
| 7.0 | 0.95 | 0.83 | < 0.001* | 0.88 [0.83 0.91] | 0.97 [0.96 0.98] | 0.61 | 0.010* |
| 8.0 | 0.94 | 0.83 | < 0.001* | 0.88 [0.84 0.91] | 0.95 [0.93 0.96] | 0.67 | 0.003* |
| 9.0 | 0.94 | 0.82 | < 0.001* | 0.89 [0.86 0.92] | 0.95 [0.93 0.96] | 0.51 | 0.035* |
| **10.0** | **0.95** | **0.84** | **< 0.001\*** | **0.89 [0.85 0.92]** | **0.96 [0.94 0.97]** | **0.68** | **0.003\*** |
| 11.0 | 0.94 | 0.82 | < 0.001* | 0.86 [0.81 0.90] | 0.94 [0.92 0.95] | 0.59 | 0.013* |
| 12.0 | 0.95 | 0.83 | < 0.001* | 0.89 [0.85 0.92] | 0.94 [0.92 0.96] | 0.62 | 0.008* |
| 13.0 | 0.95 | 0.84 | < 0.001* | 0.89 [0.85 0.92] | 0.93 [0.91 0.95] | 0.60 | 0.012* |
| 14.0 | 0.96 | 0.86 | < 0.001* | 0.86 [0.81 0.90] | 0.92 [0.89 0.94] | 0.62 | 0.008* |
| 15.0 | 0.95 | 0.83 | < 0.001* | 0.87 [0.83 0.91] | 0.92 [0.89 0.94] | 0.57 | 0.018* |
| 16.0 | 0.96 | 0.85 | < 0.001* | 0.88 [0.84 0.91] | 0.94 [0.92 0.96] | 0.61 | 0.010* |
| 17.0 | 0.95 | 0.85 | < 0.001* | 0.90 [0.86 0.92] | 0.94 [0.92 0.96] | 0.63 | 0.006* |
| 18.0 | 0.96 | 0.85 | < 0.001* | 0.89 [0.85 0.92] | 0.94 [0.91 0.95] | 0.44 | 0.080 |
| 19.0 | 0.97 | 0.88 | < 0.001* | 0.89 [0.85 0.92] | 0.94 [0.92 0.96] | 0.55 | 0.021* |
| 20.0 | 0.96 | 0.86 | < 0.001* | 0.89 [0.86 0.92] | 0.94 [0.92 0.96] | 0.53 | 0.030* |

### B. Wearable-Based Assessment of Ataxia Severity

Among the models trained on various segment lengths, the 10 s segment model was selected for detailed analysis due to its consistent performance across the validation criteria. Fig. 3a shows the cross-sectional association (*N* = 131) between clinician-evaluated BARS scores and wearable-based assessments, demonstrating a root mean square error (RMSE) of 3.6 BARS points and a strong correlation (*r* = 0.84, *p <* 0.001) [34]. Additionally, wearable-based assessments exhibited a significant correlation with clinician-scored BARS finger-to-nose subscores (*r* = 0.79, *p <* 0.001), suggesting that the estimates effectively capture both overall ataxia severity and task-specific severity.

**Fig. 3.**
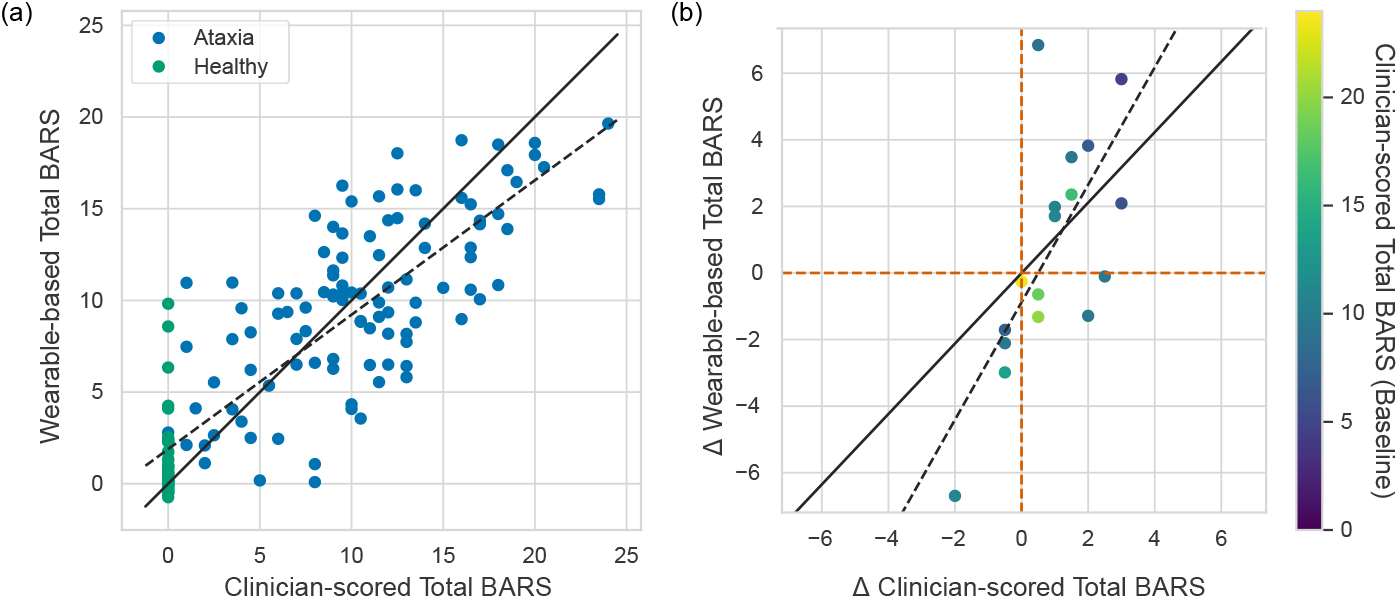
Model performance for regression task. (a) Clinician-scored total BARS vs. wearable-based total BARS from feature embeddings. Ataxia and healthy participants are represented by blue and green dots, respectively. The solid black line represents perfect estimation (***y* = *x***), while the dotted black line shows the regression line of the estimates. (b) Longitudinal changes in clinician-scored total BARS versus estimated changes in total BARS. The solid black line indicates perfect estimation (***y* = *x***), and the dotted black line represents the best linear fit. Points are color-coded according to baseline clinician-scored total BARS.

Four clinical factors were evaluated for their potential influence on wearable-based assessments: age, age-related motor development, gender, and clinical diagnosis of ataxia. To assess the effect of age, we analyzed the correlation between participant age and the model error in wearable-based assessments relative to clinician-evaluated BARS scores. A weak negative correlation was found in the overall population (*r* = −0.19, *p* = 0.019), as shown in Supplementary Fig. S2. While the results suggest that wearable-based assessments may slightly overestimate severity in younger individuals and slightly underestimate it in older individuals, model errors were symmetrically distributed around zero across the age spectrum. To further examine the impact of age-related motor development, we conducted subgroup analyses for pediatric (*<*18 years, *N* = 15) and adult (*N* = 116) participants. Neither group showed a significant correlation (pediatric: *r* = −0.23, *p* = 0.366; adult: *r* = −0.10, *p* = 0.226). Gender was also evaluated as a potential source of variability. A Mann–Whitney U test revealed no significant difference in model error between female and male participants (*p* = 0.494). To assess the model’s generalizability across heterogeneous ataxia types, we conducted a subgroup analysis based on clinical diagnosis categories listed in Table I, focusing on groups with at least seven participants. As shown in Supplementary Table S1 and Fig. S1, model errors for each subgroup were comparable to those of the overall ataxia group. While slightly higher errors were observed in participants with Ataxia-Telangiectasia and Spinocerebellar Ataxia Type 6, wearable-based assessments still demonstrated strong crosssectional correlations with BARS scores within these subgroups. Overall, these findings suggest that age, age-related motor development, gender, and clinical diagnosis are not major contributors to model error. Nevertheless, further validation in larger and more diverse populations—particularly among pediatric participants and underrepresented diagnostic groups—is warranted.

Fig. 3b presents the responsiveness of the wearable-based assessment, depicting changes in clinical scores versus changes in wearable-based assessments for participants with multiple data collection sessions (*N* = 15). A strong correlation was observed between longitudinal changes in clinical scores and the corresponding changes in wearable-based assessment (*r* = 0.68, *p* = 0.003) [34].

### C. Consistency of Ataxia Severity Assessment

Fig. 4a illustrates the agreement between wearable-based assessments derived from feature embeddings of the first half versus the second half of a finger-to-nose time-series. The color gradient represents clinician-scored motor severity (i.e., total BARS scores). The results demonstrate excellent consistency [32], with an ICC(2,1) of 0.96 (*p <* 0.001). These findings suggest that the contrastive learning algorithm reliably captures phenotypes consistently through the motor task.

**Fig. 4.**
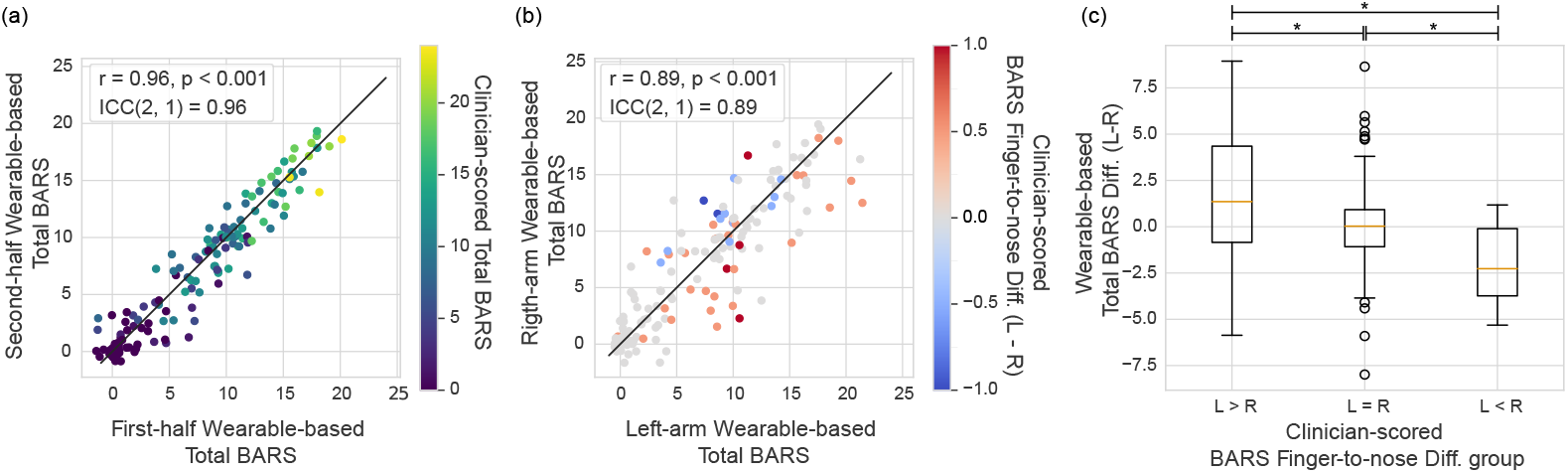
Model reliability. (a) Agreement between first-half and second-half wearable-based total BARS from a finger-to-nose task session. The points are color-coded by clinician-scored total BARS. The solid black line represents perfect agreement (***y* = *x***). (b) Agreement between left-arm and right-arm wearable-based total BARS. The color of points represents the difference in clinician-scored BARS finger-to-nose subscores between left and right arms (Left - Right). The solid black line represents perfect agreement (***y* = *x***). (c) Boxplots showing the difference in wearable-based total BARS between left and right arms (L-R), categorized by clinician-scored finger-to-nose subscores (L***>***R, L**=**R, L***<***R). Significant differences (***p <* 0.05**) are indicated with asterisks.

Fig. 4b shows the agreement between wearable-based assessments derived from the left vs. right wrists, with an ICC(2,1) of 0.89, indicating good reliability [32]. The slightly lower consistency is attributed to inherent differences in motor severity observed between the arms during the finger-to-nose task. For example, each data point in Fig. 4b is color-coded by the difference in the clinician-evaluated BARS finger-to-nose subscore (Left minus Right), where warmer colors represent greater severity in the left arm, and cooler colors indicate the opposite. Notably, most warmer-colored data points fall below the black line (*y* = *x*), suggesting that the wearable-based assessments also detected greater severity in the left arm. Likewise, cooler-colored data points are positioned above the black line, reflecting greater severity in the right arm.

To quantitatively evaluate this agreement, participants were grouped based on the difference in clinician-evaluated BARS finger-to-nose subscores: left greater than right (L*>*R, *N* = 32), equal (L=R, *N* = 109), or left less than right (L*<*R, *N* = 12). The mean differences in wearable-based total BARS estimates between left and right wrists were compared across these groups. The differences were statistically significant with a medium effect size (Kruskal–Wallis H test: *η*^2^ = 0.08, *p* = 0.001) as shown in Fig. 4c [35]. Post-hoc pairwise comparisons using the Mann-Whitney U test further supported this finding, revealing statistically significant differences in all pairs: *p* = 0.019 for L*>*R vs. L=R, *p* = 0.008 for L=R vs. L*<*R, and *p* = 0.004 for L*>*R vs. L*<*R. These preliminary results indicate that the learned feature embeddings offer the potential to capture differences in ataxia severity between arms.

### D. Detection of Ataxia

Fig. 5a shows the ROC curve for the 10 *s* model in classifying ataxia versus healthy individuals, further illustrating that the learned features encode phenotypic information associated with ataxia. The AUC-ROC was 0.95, reflecting outstanding discriminative performance [33]. Notably, for ataxia participants who scored 0 on the finger-to-nose subscore of the BARS (indicating no visually evident ataxia, *N* = 9), the wearable-based assessments differentiated them from healthy individuals with the AUC-ROC of 0.90, as shown in Fig. 5b. This preliminary finding supports the model’s potential to detect subclinical abnormalities present during the finger-to-nose task that may not be fully captured by clinical assessments.

**Fig. 5.**
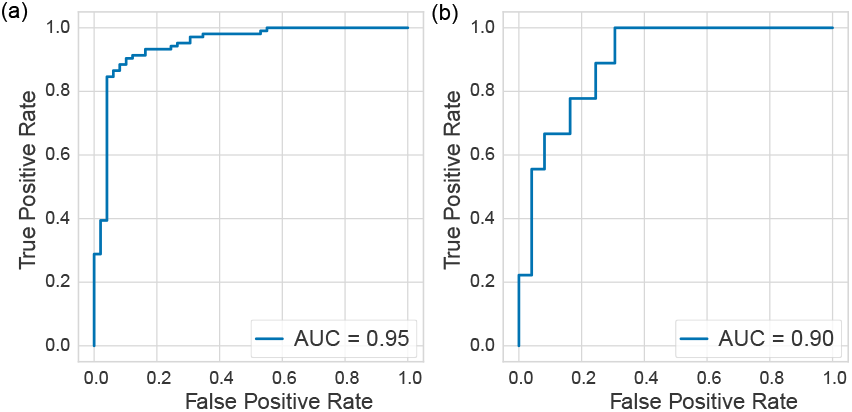
Model performance for classification task. Receiver Operating Characteristic curve illustrating the classification performance between (a) healthy individuals and those with ataxia, and (b) healthy individuals and ataxia participants who scored 0 on the finger-to-nose subscore of the BARS.

The age difference between the healthy and ataxia groups was statistically significant (Mann–Whitney U test: *U* = 839.5, *p <* 0.001). To evaluate the potential confounding effect of age, model performance was examined in a subset of participants aged ≥ 35 years. In this age-restricted subset (healthy: *N* = 16; ataxia: *N* = 73), the age difference was not statistically significant (Mann–Whitney U test: *U* = 415.5, *p* = 0.072), and the AUC-ROC remained high at 0.94. These findings suggest that age is unlikely to be a major confounding factor in the known-group validity of the model.

### E. Visualization of the Representations Learned by the Contrastive Model

Fig. 6 shows 2D-projected visualizations of the learned feature embeddings using PCA across all five dataset folds, along with a combined view. Data points are color-coded by clinician-assigned BARS scores. The first two principal components capture the majority of variance across all folds, presenting a latent manifold within the embeddings. Notably, the first principal component showed a strong correlation with clinician-assigned BARS scores, despite not using the trained regression model with the clinician-evaluated scores (Pearson’s *r* = 0.81, *p <* 0.001). This observed association across all folds highlights the embeddings’ ability to represent ataxia severity, further corroborating the results presented earlier in this section. Moreover, the similar shape of the 2D projections and the stable relationships with BARS scores support that the different models trained during each fold learn a consistent latent manifold present in the data.

**Fig. 6.**
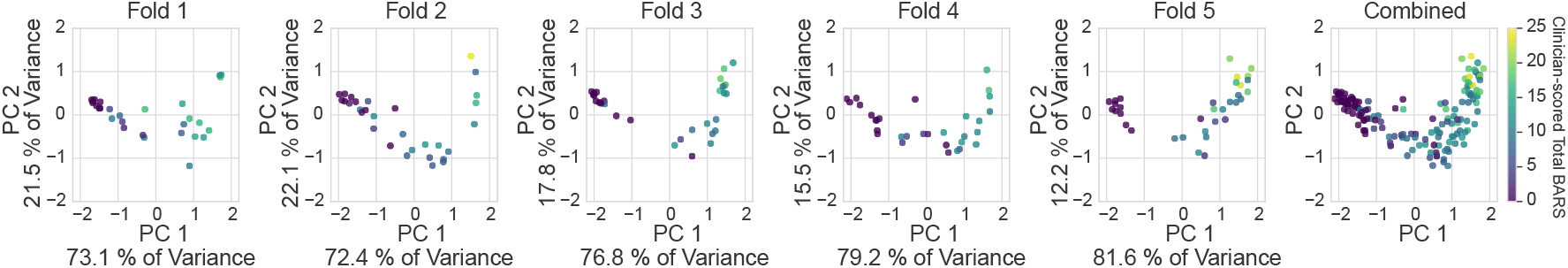
Principal Component Analysis (PCA) of the feature embeddings for five different training folds and a combined view of all folds. The plots show the first principal component (PC 1) versus the second principal component (PC 2). Each point is color-coded by the clinician-scored total BARS. The similar patterns observed across folds indicate consistency in the representation of ataxia severity, demonstrating the alignment between feature embeddings and clinician-scored severity.

## IV. Discussion

This study proposes a novel contrastive learning framework for wearable-based ataxia assessment. Notably, the learning procedure for this framework does not rely on the exact scores of clinician-rated scales. Instead, by employing a pairwise loss function, the model enhances its ability to learn subtle differences and similarities in motor severity, reducing the potential influence of biases and errors introduced by human raters or the rating scales themselves. By attaching task-specific models to the learned embeddings, we demonstrated that the embeddings encode information strongly related to the presence of ataxia and its severity. The models based on these embeddings demonstrated high reliability, sensitivity to disease progression, and an ability to differentiate subclinical signs of ataxia.

Previous studies have suggested various models for wearable-based ataxia severity assessment. For instance, Oubre *et al*. extracted manually engineered features and trained a conventional supervised machine learning model directly on total BARS scores to identify relevant features, using the same dataset as our study [12]. In contrast, our method employed representation learning with a pairwise contrastive loss to learn features, focusing on capturing a latent characteristic associated with motor severity, rather than fitting directly to clinician-evaluated scores. Oubre *et al*. reported a cross-sectional correlation between their estimates and clinical scores with Pearson’s *r* = 0.83 and a longitudinal correlation between changes in clinician-scored BARS and their estimates with *r* = 0.57. In comparison, our model achieved a cross-sectional correlation of *r* = 0.84 and a longitudinal correlation of *r* = 0.68, demonstrating improved longitudinal performance.

Gupta *et al*. extracted submovement features from wrist- and ankle-worn sensors to develop a machine-learned, wearable-based severity score independent of clinical ratings for amyotrophic lateral sclerosis, a neurodegenerative disorder affecting movement. Their approach utilized intra-subject pairwise comparisons of disease progression direction to generate severity scores, aiming to remove the reliance on subjective clinical scales. While the wearable-based severity scores demonstrated strong cross-sectional and longitudinal correlations with clinical ratings, this approach may be limited by the assumption that disease progression follows a unidirectional trajectory. This constraint could overlook fluctuating or nonlinear changes in severity, making it less applicable to conditions with variable progression patterns. In the context of ataxia, while many hereditary and neurodegenerative forms are progressive, several types (e.g., stroke-related ataxia) do not show continuous progression, limiting the direct applicability of this method to ataxia severity assessment.

We envision that the flexibility of our framework allows it to support multimodal approaches for assessing a broader range of ataxia symptoms by incorporating various types of movement time-series data from different body parts, motor tasks, or sensor modalities. For instance, the model could be extended to speech data from microphones or gait data from inertial sensors to evaluate the severity of symptoms such as slurred speech or unsteady walking in individuals with ataxia. Several prior studies have utilized multimodal data to assess ataxia severity, employing distinct manual feature extraction methods tailored to each task and sensor [14], [36]. While effective, these approaches require domain-specific knowledge and substantial effort to design features from each sensing modality based on the expected motor symptoms. In contrast, our model can offer a unified and automated approach that eliminates the need for complex, task- or sensor-specific feature engineering. By preprocessing multimodal data into a time-series format, our framework can be seamlessly applied to data from various sensors and tasks to generate learned representations. These learned representations from different modalities can be aggregated to support a more comprehensive and holistic assessment of ataxia severity. Moreover, the model has the flexibility to be expanded to incorporate relative differences from diverse sources, such as other clinical measures, ataxia subtypes, or individual longitudinal disease progression [37]. This versatility positions it as a powerful tool for capturing multiple dimensions of ataxia severity.

This study has several limitations. First, the size of longitudinal data is relatively small. Additional longitudinal data would enable a more robust evaluation of the model’s sensitivity to changes in disease progression over time. Second, the healthy and ataxia groups were not age-matched. Although an analysis using an age-restricted subset (participants aged ≥ 35 years) suggested that age is unlikely to be a major confounding factor, the lack of complete age matching remains a limitation. Future studies should aim to use age-matched groups when evaluating known-group validity. Third, although supervised contrastive learning is robust to noisy labels, it still relies on label information and thus remains partially dependent on clinician-rated scales, which are inherently subjective and often exhibit non-linear relationships with perceived motor severity. To further reduce this dependency, future work could explore self-supervised contrastive learning approaches, which have shown promise in analyzing medical time-series data [38], [39]. In addition, incorporating a wider range of clinical outcome measures—including both patient-reported and clinician-rated assessments—would help provide a more comprehensive evaluation of the model’s clinical utility. Lastly, the feature embeddings learned by our model did not have clear clinical interpretability based on visual inspection, largely due to their high dimensionality and abstract nature. While we applied PCA-based dimensionality reduction to visualize these embeddings and their relationship with clinical severity, directly interpreting them in a clinically meaningful way remains challenging and an important direction for future work. Future efforts will explore advanced interpretability techniques, such as post-hoc analysis methods [40] and attention-based architectures [41], to better understand which aspects of motor severity are captured by the learned representations and how these insights can be aligned with clinical understanding. Building on this direction, future work will also investigate how demographic factors and ataxia subtypes may influence the learned representations. These insights could guide model fine-tuning or conditioning on demographic or clinical diagnosis information to enhance sensitivity for monitoring disease progression and detecting early pathological changes.

## V. Conclusion

This study demonstrates that the proposed contrastive learning model is an effective approach for assessing ataxia severity using wrist-worn sensors during the finger-to-nose task. The model shows promising results to achieve high correlations with clinician-scored severity and demonstrating good sensitivity to changes over time. Moreover, the model exhibits strong reliability within sessions. By leveraging a pairwise loss function, the model mitigates reliance on subjective clinical scores, capturing more robust representations of disease severity. These findings highlight the potential of contrastive learning to support more objective and sensitive monitoring of ataxia progression and treatment efficacy, with clinical applications in continuous monitoring and clinical trials. Future work will focus on expanding the dataset to include more longitudinal data, refining model interpretability, and extending the framework to capture additional ataxia symptoms through multimodal data.

## Data Availability

All data produced in the present study are available upon reasonable request to the authors.

## Acknowledgments

The authors would like to thank Mary Donovan, Winnie Ching, and Nergis Khan for recruitment and data collection.

