## Supplementary Materials for "Contrastive Learning Model for Wearable-based Ataxia Assessment"

#### 1 Clinical Diagnoses and Model Performance

To examine the model's performance across heterogeneous ataxias, a subgroup analysis was conducted on clinical diagnosis groups with at least seven participants. Table S1 summarizes the root mean squared errors (RMSEs) computed for each clinical diagnosis by comparing the wearable-based assessments with clinician-scored total BARS. The mean and standard deviation of the clinician-scored BARS were provided a reference for the clinical severity of each diagnosis. In addition, Figure S1 illustrates cross-sectional associations for each diagnosis. The RMSEs observed in each subgroup are generally comparable to that of the overall ataxia group. Although Ataxia-Telangiectasia (A-T) and Spinocerebellar Ataxia Type 6 (SCA-6) show slightly higher RMSEs, the model still captures strong cross-sectional associations (Pearson's correlation: $r = 0.86$, $p = 0.013$ for SCA-6; $r = 0.75$, $p = 0.053$ for A-T), as shown in Figure S1. The correlation for A-T does not reach statistical significance, likely due to the small sample size.

Table S1: Clinical Diagnoses and Model Performance

| Clinical Diagnoses | $N$ | Clinician-scored Total BARS | RMSE | Cross-sectional correlation | |
|---|---|---|---|---|---|
| | | | | r | p |
| Overall ataxia | 87 | 10.5 ± 5.4 | 4.0 | 0.69 | < 0.001* |
| SCA-3 | 11 | 9.7 ± 3.6 | 3.2 | 0.56 | 0.073 |
| SCA-6 | 7 | 13.1 ± 5.3 | 4.4 | 0.86 | 0.013* |
| A-T | 7 | 11.4 ± 5.3 | 4.3 | 0.75 | 0.053 |
| MSA-C | 7 | 12.1 ± 3.7 | 2.9 | 0.47 | 0.285 |

RMSE: Root Mean Square Error;
SCA: Spinocerebellar ataxia;
A-T: Ataxia-Telangiectasia;
MSA-C: Multiple system atrophy, cerebellar-type

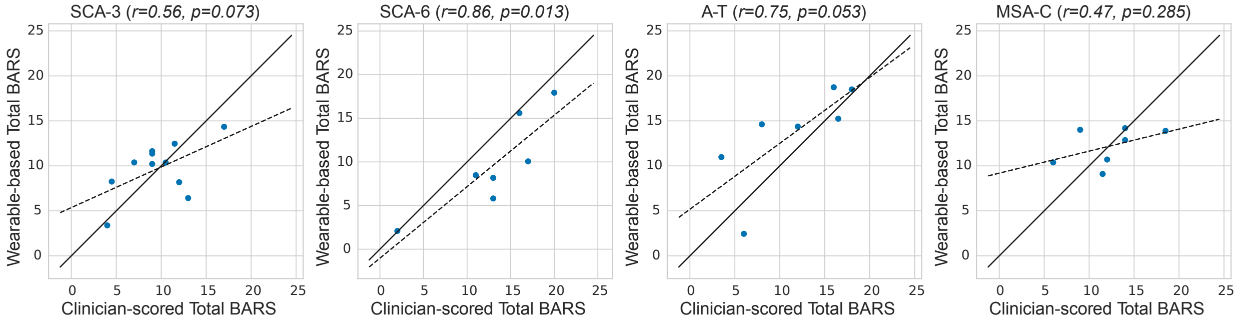


Figure S1: Clinician-scored total BARS vs. wearable-based total BARS for participants diagnosed with SCA-3, SCA-6, A-T, and MSA-C. The solid black line represents perfect agreement ($y = x$), while the dotted black line indicates the least-squares fit with the model estimates.

### 2 Age Analysis

To investigate the potential impact of age-related changes in movements on model estimates, the relationship between age and model error was analyzed as shown in Figure S2. A weak negative correlation was observed in the overall population ($r = -0.19$, $p = 0.019$), indicating slightly lower model errors for older individuals. However, as shown in Figure S2, model errors were symmetrically distributed around zero across the age range, indicating that age may not be a dominant source of model bias. To better understand whether this relationship reflects age-related motor development, particularly in pediatric populations, subgroup analyses were conducted separately for pediatric ($<$ 18 years, $n = 15$) and adult ($n = 116$) participants. However, no statistically significant correlation was found in either the pediatric ($r = -0.23$, $p = 0.366$) or adult ($r = -0.10$, $p = 0.226$) group. These findings suggest that age-related motor development is unlikely to be a major contributor to model error, although confirmation in larger pediatric samples is necessary.

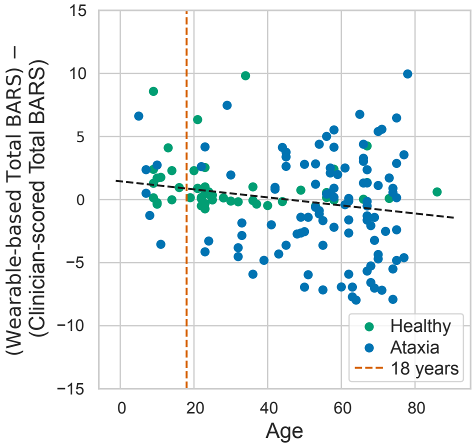


Figure S2: Age vs. model error. Healthy and ataxia participants are represented by green and blue dots, respectively. The red dotted line indicates 18 years of age.
